# Quality of life of Type II Diabetic patients

**DOI:** 10.1101/2024.11.07.24316896

**Authors:** Sulochana Ghimire, Gita Neupane, Chanda Sah, Mahesh Raj Ghimire, Binita Soti

## Abstract

**Objective:** Diabetes mellitus poses significant challenges to individuals’ well-being, affecting various aspects of their lives beyond the physical symptoms of the disease. Understanding the multidimensional aspects of QoL among diabetic patients is crucial for providing holistic healthcare interventions and improving overall health outcomes. The study aims to evaluate the quality of life of Type II Diabetes Mellitus patients.

**Research Design and Methods:** Descriptive cross-sectional study was conducted among 334 diagnosed cases of Type 2 Diabetes Mellitus for more than or equal to 6 months attending the outpatient department of UCMS-TH. Non-probability purposive sampling technique was used to select samples for the study. The WHOQOL-BREF questionnaire was used to measure QoL. Data were analyzed with descriptive statistics (frequency, percentage, mean, standard deviation) and inferential statistics (t-tests or one way ANOVA) to explore associations between QoL domains and sociodemographic characteristics.

**Results:** More than half (56.6%) of the respondents were between the age group of 41-60 years with mean age of 58.42. Highest mean score ± SD was found in social domain (60.77 ± 13.83) followed by environmental domain (56.05 ± 10.38) and psychological domain (55.67 ±8.44) with least mean domain score in physical domain (49.99 ± 14.53). The results show that diabetic patients, particularly women and those with comorbid conditions, report lower quality of life in all domains. Additionally, no significant association was found between having a family history of diabetes and quality of life. There was high positive correlation between physical and environmental domain of quality of life (r = 0.70, p < 0.001).

**Conclusion:** Comprehensive management strategies focusing on all dimensions of health is necessary to improve the quality of life of patients with Diabetes Mellitus.

## Introduction

### Background of the study

Diabetes Mellitus is a chronic disease that represents a significant global health challenge and is classified among the top four priority noncommunicable disease [1]. An estimated 3 in 4 people are having Diabetes Mellitus in low and middle-income countries [2, 3]. Prevalence of prediabetes and diabetes rose gradually with advancing age. Diabetes Mellitus prevalence varied significantly across Nepal’s provinces, ranging from as low as 2% in Province 6 to as high as 10% in Province 3 and 4 [4]. Diabetes Mellitus complications include macrovascular (coronary artery disease, peripheral arterial disease, stroke) and microvascular (nephropathy, neuropathy, retinopathy) [5]. Diabetes Mellitus not only affect physical health but also affect social and mental including psychological well-being of people living with it. The psychosocial issues frequently experienced by individuals with diabetes often have substantial adverse effects on their well-being [6]. Individuals diagnosed with type 2 diabetes mellitus (T2DM) encounter notable obstacles during their treatment journey, potentially hindering effective disease control. Effective management of the condition can alleviate symptoms, enhance glycemic control, prevent complications and minimize hospital readmissions and mortality [7]. A qualitative study showed that certain adults e.g., adults aged >45 years of age with type 2 diabetes mellitus encountered both physical and psychological ailments. Managing life with type 2 Diabetes Mellitus was impacted by factors including family support, inadequate adherence to treatment protocols, and availability of information, education, and communication resources [8]. Increasing prevalence of Diabetes Mellitus will lead to a higher incidence of chronic and acute illnesses within the general population, resulting in significant implications for quality of life, healthcare service demand, and economic expenditures [9]. Diabetes therapy, like insulin use, impacts quality of life both positively by lowering high blood sugar and negatively by raising low blood sugar. The psychosocial burden also affects self-care and raises the risk of long-term complications that reduce QoL [10]. The WHO defines Quality of Life as an individual’s view of their position in life, shaped by cultural, value systems, goals, expectations, standards, and concerns [11]. All members of the interprofessional healthcare team should acknowledge the importance of quality of life [12]. Diabetes Mellitus significantly impacts on QoL, affecting physical health, emotional well-being, social interactions, and finances. Effective management strategies, including education, support, and access to healthcare resources, are essential for improving QoL for individuals living with Diabetes Mellitus. Understanding the quality of life of patients living with type 2 diabetes Mellitus is essential for formulating strategies to support patient-centered care, optimize treatment modalities, and promote overall well-being. This study aimed to assess the quality of life of people with a diagnosed Type 2 Diabetes Mellitus from a tertiary care center of Nepal.

## Materials and Methods

### Study Setting

A Cross sectional descriptive study was conducted to find out the quality of life of diabetic patients. The study was conducted in Universal College of Medical Sciences, Teaching Hospital (UCMS-TH) Rupandehi, Nepal which is 750 bedded well equipped multi-specialty hospital. Participants were patients who were diagnosed with Type II Diabetes Mellitus for at least 6 months, attending outpatient department of UCMS-TH, who consented to the study and were available to provide information during data collection.

### Sampling

Sample size was determined by using Cochran’s formula by taking 95% confidence level and the prevalence of good quality of life of diabetic patients (P) = 68% from a study conducted in rural south India [13]. Based on calculation 334 diabetic patients were involved in this study by using non-probability purposive sampling technique and respondents were interviewed in first come first basis who visited the OPD.

### Research Instrument

For data collection structured interview questionnaire was developed which consisted of following 2 parts: First part socio-demographic information questionnaire contains age, gender, marital status, education, occupation, family history of diabetes, years of diagnosis of diabetes, modalities of treatment and co-morbidities related questions.

Second part WHOQOL-BREF is a 26-item questionnaire that measures an individual’s quality of life. The WHOQOL BREF assesses quality of life across four domains: physical health, psychological well-being, social relationships and environment. It also includes two separate questions about overall quality of life and health perception. Each individual item of WHOQOL-BREF is scored from 1 to 5 on a response scale [14]. Domain scores are scaled in a positive direction (i. e. higher scores denote higher quality of life). Three items (i.e., item no.3, item no.4 & item no.26) are phrased negatively so before calculating raw score, 3 items were reversed scored. Mean score in each domain is multiplied by 4 to align with WHOQOL-100 scores, then converted to a 0-100 scale, where 100 indicates the highest and 0 the lowest quality of life, as per the WHO scoring manual. The WHOQOL-BREF had a reliability of 0.896, as measured by Cronbach’s alpha coefficient and test–retest analysis [15]. Permission was taken from WHO reproduce, reprint and/or translate WHOQOL Questionnaire. English version questionnaire was translated to Nepali language and was back translated before data collection. Pretesting of the instrument was done on 10% (i. e. 33 respondents) of total sample size.

### Ethical Approval

Ethical clearance was taken from Institutional Review Committee of Universal College of Medical Science (UCMS/IRC/030/23). Administrative approval for data collection was provided by Medical Superintendent of UCMS-TH. Written informed consent was secured from each respondent after explaining the study’s objective. Respondents were assured for the confidentiality of the information. Collected data was checked immediately for completion and was coded, edited, classified, entered and cleaned. Researcher herself collected the data within six months periods (01^st^ October 2023 to 29^th^ March 2024).

### Statistical Analysis

The reliability of pretest measured by Cronbach’s alpha was 0.880. The Kolmogorov-Smirnov normality test scores showed normal distribution with P Value 0.068. Data analysis was done by using SPSS version 20. Association between QoL domains and sociodemographic characteristics were tested using t – test or one way ANOVA.

## Results

A total 334 respondents participated in the study. Socio-demographic related information of the respondents is shown in table 1. More than half (56.6%) of the respondents were between the age group of 41-60 years with mean age of 58.42 ± 10.34 SD. Twenty nine percent of the respondents perceived their overall good quality of life as and 21.9% of the respondents were satisfied about their health assessed by 1^st^ and 2^nd^ general question of WHO QoL BREF Questionnaire respectively.

**Table 1:** Socio-demographic characteristics of respondents.

| N= 334 |  |  |
| --- | --- | --- |
| Variables | Frequency | Percentage |
| **Age (in completed years)** |  |  |
| 21- 40 | 13 | 3.9 |
| 41- 60 | 189 | 56.6 |
| >60 | 132 | 39.5 |
| **Mean ± SD (58.42 ± 10.34)** |  |  |
| **Gender** |  |  |
| Male | 182 | 54.5 |
| Female | 152 | 45.5 |
| **Marital Status** |  |  |
| Married | 287 | 85.9 |
| Unmarried | 11 | 3.3 |
| Divorced | 36 | 10.8 |
| **Education** |  |  |
| Illiterate | 125 | 37.5 |
| Literate | 209 | 62.5 |
| **Occupation** |  |  |
| Business | 76 | 22.8 |
| Service | 86 | 25.7 |
| Agriculture | 55 | 16.5 |
| Retired | 15 | 4.5 |
| Homemaker | 102 | 30.5 |
| **Family History of Diabetes Mellitus** |  |  |
| Yes | 197 | 59.0 |
| No | 137 | 41.0 |
| **Duration of Diagnosis of Diabetes Mellitus** |  |  |
| ≤ 5 years | 169 | 50.6 |
| 6-10 years | 124 | 37.1 |
| > 10 years | 41 | 12.3 |
| **Treatment used** |  |  |
| Diet + Oral Agents | 278 | 83.2 |
| Diet + Insulin | 45 | 13.5 |
| Diet + Oral agents + Insulin | 11 | 3.3 |
| **Comorbidities** |  |  |
| Yes | 179 | 53.6 |
| No | 155 | 46.4 |

The four domains namely physical, psychological, social and environmental score denote an individual’s perception of quality of life in each particular domain and was analyzed according to WHO QoL BREF questionnaire. Transformed scores were calculated from raw score to compare with WHOQOL – 100 score as shown in table 2. Highest mean score ± SD was found in social domain (60.77 ± 13.83) followed by environmental domain (56.05 ± 10.38) and psychological domain (55.67 ±8.44) with least mean domain score in physical domain (49.99 ± 14.53).

**Table 2:** WHO-BREF QOL Domain score of respondents.

**N=334**
| WHO-BREF QOL Domain | Raw Score |  | Transformed score (0-100) |  |
| --- | --- | --- | --- | --- |
| | Mean score ± SD | Range | Mean score ± SD | Range |
| Physical Domain | $20.29 \pm 4.07$ | 12 – 28 | $49.99 \pm 14.53$ | 19-75 |
| Psychological Domain | $19.37 \pm 2.00$ | 14 – 24 | $55.67 \pm 8.44$ | 31-75 |
| Social Domain | $10.30 \pm 1.62$ | 6 – 15 | $60.77 \pm 13.83$ | 25-100 |
| Environmental Domain | $25.45 \pm 3.27$ | 16 - 34 | $56.05 \pm 10.38$ | 25-81 |

In age group statistically significant differences was found in four domains of QoL. For physical and psychological domain, mean score for those of more than 60 years of age group was lower as compared to 21-40 or 41-60 years of age groups. For social and environmental domain, the mean score of more than 60 years of age group was lower as compared to mean score of 41-60 years of age group.

There was statistically significant difference between gender in four domains of QoL. In all domains, mean score for male was higher than female. In terms of marital status, there was statistically significant differences in physical and environmental domain. For physical and environmental domain, the mean score of divorced was significantly lower as compared to unmarried group. Likewise, there was statistically significant difference between educational status in all four domains of Quality of Life. In all domains, mean score for literate group of respondents was higher than illiterate group of respondents.

In terms of duration of diagnosis of Diabetes Mellitus there was statistically significant differences in four domains of QoL. For physical and environmental domain, mean score of respondents whose Diabetes Mellitus was diagnosed in ≤ 5 years was higher as compared to mean score of respondents whose duration of Diabetes Mellitus was 6-10 or >10 years. For psychological domain, the mean score of respondents whose Diabetes Mellitus was diagnosed in ≤ 5 years was higher as compared to mean score of respondents whose duration of Diabetes Mellitus was 6-10 years. For social domain, the mean score of respondents whose Diabetes Mellitus was diagnosed in ≤ 5 years was higher as compared to >10 years of duration of illness.

Similarly, in treatment used, there was statistically significant differences in psychological and environmental domain. For psychological domain, the mean score of respondents using diet and oral hypoglycemic agents for managing Diabetes Mellitus have higher mean score as compared to the mean score of groups of respondents using diet and insulin or diet, oral hypoglycemic agents and insulin for managing Diabetes Mellitus.

There were statistically significant differences between the presence of comorbidities in all four domains of quality of life. In all domains, mean score of the respondents without any comorbidities was higher as compared to the mean score of the respondents having comorbidities as shown in table 3.

**Table 3:**
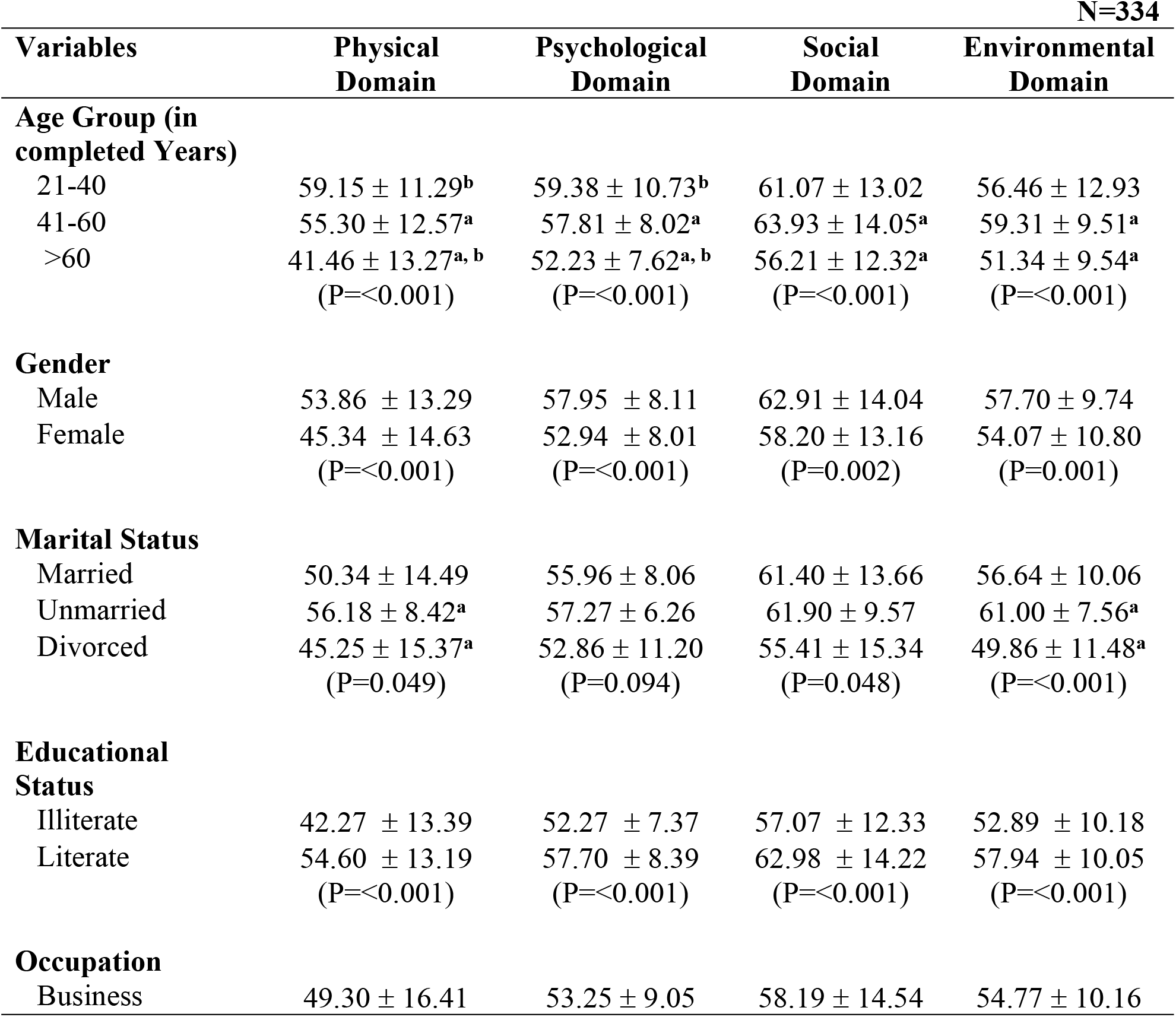
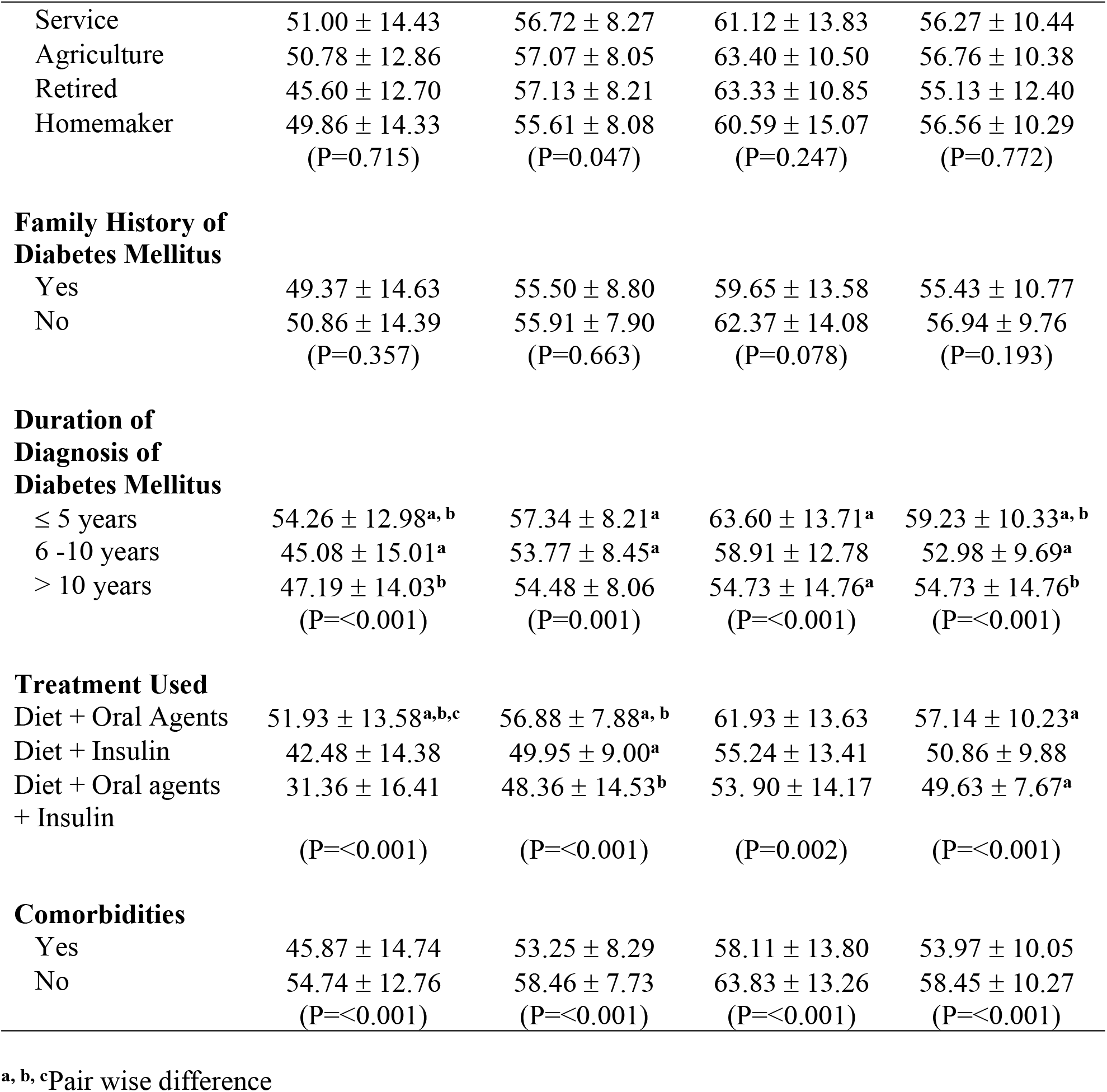
Association between WHO QoL BREF domains and Sociodemographic Characteristics.

**N=334**
| Variables | Physical Domain | Psychological Domain | Social Domain | Environmental Domain |
| --- | --- | --- | --- | --- |
| **Age Group (in completed Years)** |  |  |  |  |
| 21-40 | $59.15 \pm 11.29^b$ | $59.38 \pm 10.73^b$ | $61.07 \pm 13.02$ | $56.46 \pm 12.93$ |
| 41-60 | $55.30 \pm 12.57^a$ | $57.81 \pm 8.02^a$ | $63.93 \pm 14.05^a$ | $59.31 \pm 9.51^a$ |
| >60 | $41.46 \pm 13.27^{a, b}$<br>($P=<0.001$) | $52.23 \pm 7.62^{a, b}$<br>($P=<0.001$) | $56.21 \pm 12.32^a$<br>($P=<0.001$) | $51.34 \pm 9.54^a$<br>($P=<0.001$) |
| **Gender** |  |  |  |  |
| Male | $53.86 \pm 13.29$ | $57.95 \pm 8.11$ | $62.91 \pm 14.04$ | $57.70 \pm 9.74$ |
| Female | $45.34 \pm 14.63$<br>($P=<0.001$) | $52.94 \pm 8.01$<br>($P=<0.001$) | $58.20 \pm 13.16$<br>($P=0.002$) | $54.07 \pm 10.80$<br>($P=0.001$) |
| **Marital Status** |  |  |  |  |
| Married | $50.34 \pm 14.49$ | $55.96 \pm 8.06$ | $61.40 \pm 13.66$ | $56.64 \pm 10.06$ |
| Unmarried | $56.18 \pm 8.42^a$ | $57.27 \pm 6.26$ | $61.90 \pm 9.57$ | $61.00 \pm 7.56^a$ |
| Divorced | $45.25 \pm 15.37^a$<br>($P=0.049$) | $52.86 \pm 11.20$<br>($P=0.094$) | $55.41 \pm 15.34$<br>($P=0.048$) | $49.86 \pm 11.48^a$<br>($P=<0.001$) |
| **Educational Status** |  |  |  |  |
| Illiterate | $42.27 \pm 13.39$ | $52.27 \pm 7.37$ | $57.07 \pm 12.33$ | $52.89 \pm 10.18$ |
| Literate | $54.60 \pm 13.19$<br>($P=<0.001$) | $57.70 \pm 8.39$<br>($P=<0.001$) | $62.98 \pm 14.22$<br>($P=<0.001$) | $57.94 \pm 10.05$<br>($P=<0.001$) |
| **Occupation** |  |  |  |  |
| Business | $49.30 \pm 16.41$ | $53.25 \pm 9.05$ | $58.19 \pm 14.54$ | $54.77 \pm 10.16$ |
| Service | 51.00 ± 14.43 | 56.72 ± 8.27 | 61.12 ± 13.83 | 56.27 ± 10.44 |
| Agriculture | 50.78 ± 12.86 | 57.07 ± 8.05 | 63.40 ± 10.50 | 56.76 ± 10.38 |
| Retired | 45.60 ± 12.70 | 57.13 ± 8.21 | 63.33 ± 10.85 | 55.13 ± 12.40 |
| Homemaker | 49.86 ± 14.33<br>(P=0.715) | 55.61 ± 8.08<br>(P=0.047) | 60.59 ± 15.07<br>(P=0.247) | 56.56 ± 10.29<br>(P=0.772) |
| **Family History of Diabetes Mellitus** |  |  |  |  |
| Yes | 49.37 ± 14.63 | 55.50 ± 8.80 | 59.65 ± 13.58 | 55.43 ± 10.77 |
| No | 50.86 ± 14.39<br>(P=0.357) | 55.91 ± 7.90<br>(P=0.663) | 62.37 ± 14.08<br>(P=0.078) | 56.94 ± 9.76<br>(P=0.193) |
| **Duration of Diagnosis of Diabetes Mellitus** |  |  |  |  |
| ≤ 5 years | 54.26 ± 12.98<sup>a, b</sup> | 57.34 ± 8.21<sup>a</sup> | 63.60 ± 13.71<sup>a</sup> | 59.23 ± 10.33<sup>a, b</sup> |
| 6 -10 years | 45.08 ± 15.01<sup>a</sup> | 53.77 ± 8.45<sup>a</sup> | 58.91 ± 12.78 | 52.98 ± 9.69<sup>a</sup> |
| > 10 years | 47.19 ± 14.03<sup>b</sup><br>(P=<0.001) | 54.48 ± 8.06<br>(P=0.001) | 54.73 ± 14.76<sup>a</sup><br>(P=<0.001) | 54.73 ± 14.76<sup>b</sup><br>(P=<0.001) |
| **Treatment Used** |  |  |  |  |
| Diet + Oral Agents | 51.93 ± 13.58<sup>a, b, c</sup> | 56.88 ± 7.88<sup>a, b</sup> | 61.93 ± 13.63 | 57.14 ± 10.23<sup>a</sup> |
| Diet + Insulin | 42.48 ± 14.38 | 49.95 ± 9.00<sup>a</sup> | 55.24 ± 13.41 | 50.86 ± 9.88 |
| Diet + Oral agents + Insulin | 31.36 ± 16.41<br>(P=<0.001) | 48.36 ± 14.53<sup>b</sup><br>(P=<0.001) | 53.90 ± 14.17<br>(P=0.002) | 49.63 ± 7.67<sup>a</sup><br>(P=<0.001) |
| **Comorbidities** |  |  |  |  |
| Yes | 45.87 ± 14.74 | 53.25 ± 8.29 | 58.11 ± 13.80 | 53.97 ± 10.05 |
| No | 54.74 ± 12.76<br>(P=<0.001) | 58.46 ± 7.73<br>(P=<0.001) | 63.83 ± 13.26<br>(P=<0.001) | 58.45 ± 10.27<br>(P=<0.001) |

<sup>a, b, c</sup>Pair wise difference

Cronbach’s alpha coefficient was used to assess the internal consistency of the WHO QoL BREF scale and its four domains. Table 4 shows significant correlations between all domains (P < 0.05). A moderate positive correlation was found between the physical and psychological domains (r = 0.63, p < 0.001), while a strong positive correlation was observed between the physical and environmental domains (r = 0.70, p < 0.001).

**Table 4:**
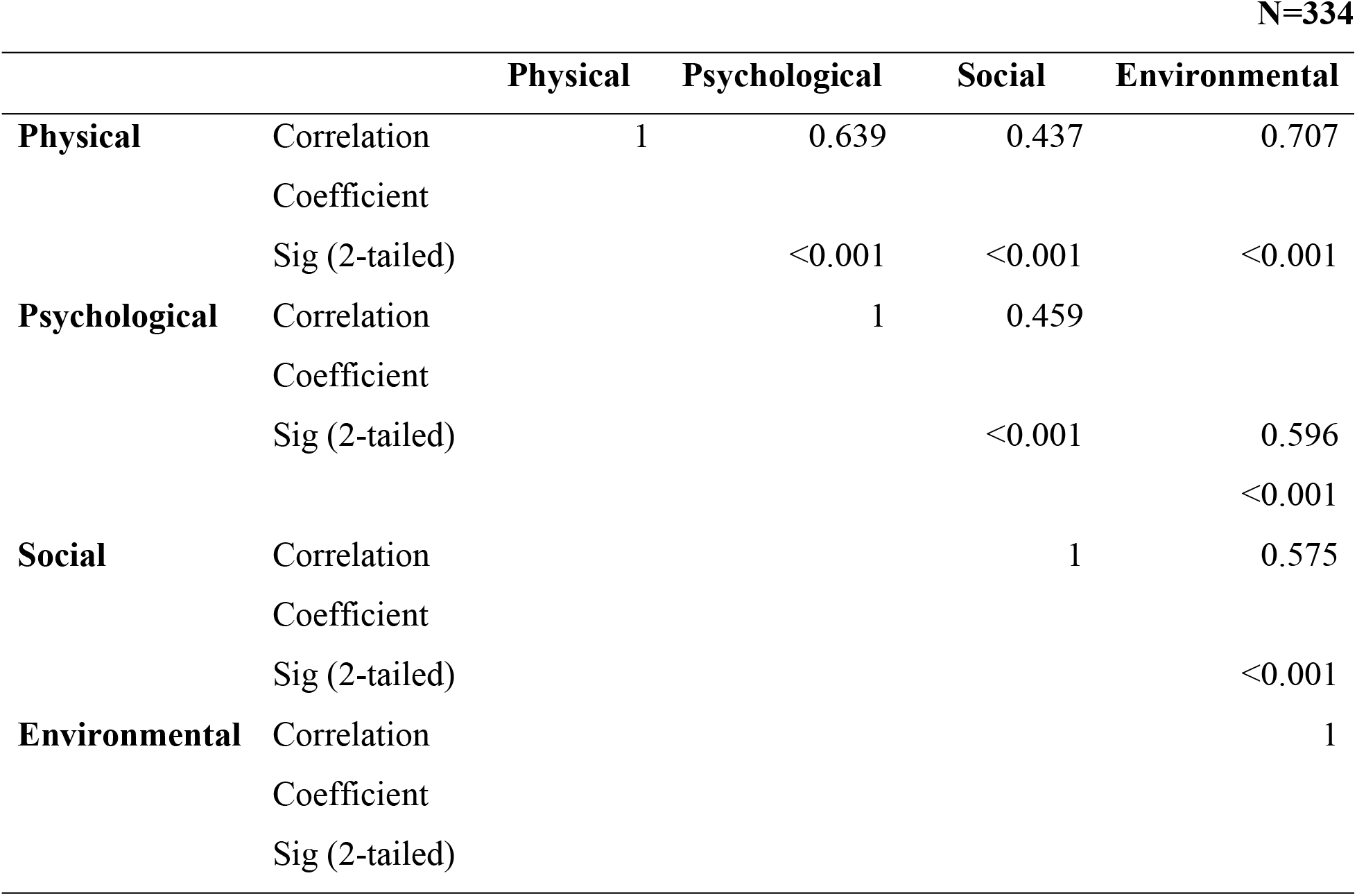
Correlation coefficient in four domains of WHO QoL BREF.

## Discussion

Prevalence of diabetes has significantly risen in both developed and developing countries over the past four decades and is largely attributed to an abundance of food, shifts in dietary habits, and a decrease in physical activity [16]. Diabetes Mellitus significantly affects individual’s quality of life making it essential to address quality of life aspects in diabetes management to improve overall outcomes.

In present study majority (56.6%) of the respondents were between the age group of 41-60 years. This finding is consistent with the study conducted in India [17] in which majority (62.13%) of the respondents were between the age group of 41-60 years. The distribution of gender (male: 54.5% and female: 45.5%) in present study is consistent with the study of India [18] and Bangladesh [19], where respondents were equally distributed between male and female. The reason of this similar findings might be because the demographic profiles of Nepal, India, and Bangladesh share significant similarities due to their geographical proximity, cultural connections, and comparable socio-economic conditions. These factors often lead to parallel patterns in population characteristics, including gender distribution.

The mean QoL scores was highest in social domain followed by environmental domain, psychological domain and physical domain. This finding is similar with the study conducted in Nepal [20]. While discussing the reasons for low quality of life scores in the physical domain among diabetes patients, it’s important to explore the various factors that contribute to physical challenges and limitations experienced by these individuals. Mental health issues can manifest physically by decreasing energy levels, increasing fatigue, and reducing the overall willingness to participate in physical activities, which are crucial for maintaining physical health and well-being. Strict dietary restrictions necessary for diabetes management can sometimes lead to nutritional deficiencies, affecting physical energy levels and overall physical health. The fear of experiencing hypoglycemia during exercise may discourage diabetes patients from participating in physical activities, leading to a sedentary lifestyle and lower quality of life in the physical domain.

In present study there was statistically significant difference between gender in all four domains of Quality of Life. This finding is consistent with the study conducted in India [13,17, 19]. The observed lower quality of life scores among females might be related to various challenges faced by females like economic disparities between male and females that might lead to delay in health seeking behavior. Furthermore, cultural roles and gender roles might place additional pressure on females, which could adversely affect their quality of life.

There was no statistically significant difference between family history of Diabetes Mellitus and all four domains of Quality of Life. This find is supported by the study conducted in eastern India [17]. The absence of a significant difference suggests that having a family history of diabetes does not directly influence how patients perceive their quality of life in four domains (e.g., physical, psychological, social relationships, and environment). This implies that variou factors such as access to healthcare, individual health behaviors, socioeconomic status, education, and support systems might have a more pronounced effect on quality of life than family history. These factors could overshadow the potential influence of family history on QoL domains.

In present study there were statistically significant differences between presence of comorbidities in all four domains of quality of life. Comorbid conditions can impose greater restrictions on daily activities, which may explain the lower quality of life scores among those with comorbidities, compared to those without such conditions who face fewer limitations. Furthermore, the reduced complexity of disease management among patients without comorbid conditions may lead to a lower healthcare burden and improved quality of life, as managing diabetes alone is less demanding than managing multiple chronic conditions simultaneously.

## Conclusion

Based on the findings of the study, it was concluded that Diabetes Mellitus places huge impact in QoL. The findings demonstrate that diabetic patients, especially females and those with comorbidities, experience lower QoL across all domains. Furthermore, no significant difference was observed between family history of diabetes and QoL, suggesting that factors beyond genetic predisposition, such as disease management and psychosocial support, may have more influential role in determining overall quality of life. The results underscore the need for a more holistic approach to diabetes care, focusing not only on glycemic control but also on addressing psychological support, lifestyle modifications, and management of comorbid conditions to improve patients’ quality of life.

## Data Availability

All relevant data are within the manuscript and its Supporting Information files.

## Acknowledgements

Authors would like to extend their sincere thanks to WHO for granting permission to use the WHOQOL-BREF scale in this study. WHOQOL-BREF provided comprehensive tool for assessing QoL among diabetic patients, which was essential for the success of this research. We would also like to thank Mr. Pradeep Chhetri and Mr. Shakti Shrestha whose statistical guidance and expertise were invaluable throughout the study. And lastly, authors express their deepest gratitude to all the respondents who participated in study. Respondents’ willingness to share their experiences and insights were invaluable to our understanding of the quality of life among diabetic patients.

## References

1. Petersmann A, Nauck M, Müller-Wieland D, Kerner W, Müller UA, Landgraf R, Freckmann G, Heinemann L. Definition, classification and diagnostics of diabetes mellitus. Journal of Laboratory Medicine. 2018 Jun 27;42(3):73–9.

2. Chen L, Magliano DJ, Zimmet PZ. The worldwide epidemiology of type 2 diabetes mellitus—present and future perspectives. Nature reviews endocrinology. 2012 Apr;8(4):228–36.

3. https://diabetesatlas.org/

4. Shrestha N, Mishra SR, Ghimire S, Gyawali B, Mehata S. Burden of diabetes and prediabetes in Nepal: a systematic review and meta-analysis. Diabetes Therapy. 2020 Sep;11:1935–46.

5. Fowler MJ. Microvascular and macrovascular complications of diabetes. Clinical diabetes. 2008 Apr 1;26(2):77–82.

6. Kalra S, Jena BN, Yeravdekar R. Emotional and psychological needs of people with diabetes. Indian journal of endocrinology and metabolism. 2018 Sep 1;22(5):696–704.

7. Nikpour S, Mehrdad N, Sanjari M, Aalaa M, Heshmat R, Mafinejad MK, Larijani B, Nomali M, Ghezeljeh TN. Challenges of type 2 diabetes mellitus management from the perspective of patients: Conventional content analysis. Interactive Journal of Medical Research. 2022 Oct 27;11(2):e41933.

8. Mwila KF, Bwembya PA, Jacobs C. Experiences and challenges of adults living with type 2 diabetes mellitus presenting at the University Teaching Hospital in Lusaka, Zambia. BMJ Open Diabetes Research and Care. 2019 Nov 1;7(1):e000497.

9. Harding JL, Pavkov ME, Magliano DJ, Shaw JE, Gregg EW. Global trends in diabetes complications: a review of current evidence. Diabetologia. 2019 Jan;62:3–16.

10. Rubin RR, Peyrot M. Quality of life and diabetes. Diabetes/metabolism research and reviews. 1999 May;15(3):205–18 Deshpande AD, Harris-Hayes M, Schootman M. Epidemiology of diabetes and diabetes-related complications. Physical therapy. 2008 Nov 1;88(11):1254–64.

11. Harper A, Orley J. WHOQOL-BREF: Introduction, administration, scoring and generic version of the assessment. Programme on Mental Health [Internet]. Geneva: World Health Organization. 1996.

12. Teoli D, Bhardwaj A. Definition/Introduction. Patient Self-Determination Act. 2020.

13. Manjunath K, Christopher P, Gopichandran V, Rakesh PS, George K, Prasad JH. Quality of life of a patient with type 2 diabetes: A cross-sectional study in Rural South India. Journal of family medicine and primary care. 2014 Oct;3(4):396.

14. Ilić I, Šipetić-Grujičić S, Grujičić J, Živanović Mačužić I, Kocić S, Ilić M. Psychometric properties of the world health organization’s quality of life (WHOQOL-BREF) questionnaire in medical students. Medicina. 2019 Dec 4;55(12):772.

15. World Health Organization. Programme on mental health: WHOQOL user manual. World Health Organization; 1998.

16. Trikkalinou A, Papazafiropoulou AK, Melidonis A. Type 2 diabetes and quality of life. World journal of diabetes. 2017 Apr 4;8(4):120.

17. Sahoo SS, Sahoo JR, Taywade M, Patro BK. Quality of life and its determinants among ambulatory diabetic patients attending NCD prevention clinic: A cross sectional study from Eastern India. Clinical Epidemiology and Global Health. 2023 May 1;21:101275.

18. Patil S, Patil Y, Patil SK. Assessment of quality of life in type 2 diabetes mellitus patients using World Health Organization quality of life-BREF questionnaire and appraisal of diabetes scale-a cross-sectional study. Italian Journal of Medicine. 2021 Oct 5;15(3).

19. Amin MF, Bhowmik B, Rouf R, Khan MI, Tasnim SA, Afsana F, Sharmin R, Hossain KN, Khan MA, Amin SM, Khan MS. Assessment of quality of life and its determinants in type-2 diabetes patients using the WHOQOL-BREF instrument in Bangladesh. BMC Endocrine Disorders. 2022 Jun 18;22(1):162.

20. Mishra SR, Sharma A, Bhandari PM, Bhochhibhoya S, Thapa K. Depression and health-related quality of life among patients with type 2 diabetes mellitus: a cross-sectional study in Nepal. PloS one. 2015 Nov 23;10(11):e0141385.

